# Genetic and neurophysiological biomarkers of neuroplasticity inform post-stroke language recovery

**DOI:** 10.1101/2021.07.28.21260995

**Authors:** Haley C. Dresang, Denise Y. Harvey, Priyanka P. Shah-Basak, Laura DeLoretta, Rachel Wurzman, Shreya Y. Parchure, Daniela Sacchetti, Olufunsho Faseyitan, Falk W. Lohoff, Roy H. Hamilton

## Abstract

**Background:** There is high variability in post-stroke aphasia severity and predicting recovery remains imprecise. Standard prognostics do not include neurophysiological indicators or genetic biomarkers of neuroplasticity, which may be critical sources of variability.

**Objective:** To evaluate whether a common polymorphism (Val^66^Met) in the gene for brain-derived neurotrophic factor (BDNF) contributes to variability in post-stroke language recovery, and to assess whether BDNF polymorphism interacts with neurophysiological indicators of neuroplasticity to improve estimates of aphasia severity.

**Methods:** Saliva samples and motor-evoked potentials (MEPs) were collected from participants with chronic aphasia subsequent to left-hemisphere stroke. MEPs were collected prior to continuous theta burst stimulation (cTBS; index for cortical excitability) and 10 minutes following cTBS (index for stimulation-induced neuroplasticity) to the left primary motor cortex. Analyses assessed the extent to which BDNF polymorphism interacted with cortical excitability and stimulation-induced neuroplasticity to predict aphasia severity beyond established predictors.

**Results:** Val^66^Val carriers showed less aphasia severity than Met allele carriers, after controlling for lesion volume and time post-stroke. Furthermore, Val^66^Val carriers showed expected effects of age on aphasia severity, and positive associations between both cortical excitability and stimulation-induced neuroplasticity and severity. In contrast, Met allele carriers showed weaker effects of age and negative associations between cortical excitability, stimulation-induced neuroplasticity and aphasia severity.

**Conclusions:** Neurophysiological indicators and genetic biomarkers of neuroplasticity improved ability to predict aphasia severity. Furthermore, BDNF polymorphism interacted with cortical excitability and stimulation-induced neuroplasticity to improve predictions. These findings provide novel insights into mechanisms of variability in stroke recovery and may improve aphasia prognostics.

## Background

Post-stroke aphasia severity and potential for language recovery are highly variable and difficult to predict. Common prognostic indicators of aphasia severity include inter-related variables such as neurological (e.g., lesion size, location) and demographic (e.g., age, time post-stroke) profiles[1]. However, standard prognostics do not include neurophysiological indicators or biomarkers of neuroplasticity. These are critical variables to examine because neuroplastic reorganization is necessary for biological and behavioral recovery after stroke or other forms of brain injury[2–8].

This research examined the hypothesis that individual differences in biomarkers of neuroplasticity improve predictions of aphasia severity. In particular, we examined whether individual differences in neuroplasticity – including a genetic biomarker and neurophysiological indicators of cortical excitability and neuroplasticity – improved ability to predict aphasia severity in subjects with chronic post-stroke aphasia. Below we summarize evidence that indicates each of these factors may account for variance in post-stroke aphasia.

First, we examined ***brain-derived neurotrophic factor (BDNF)***, which is a protein encoded by the BDNF gene. BDNF is a neurotrophin critical for neural repair and plasticity. It exhibits activity-dependent release at synapses[9] and modulates long-term potentiation[10] and long-term depression processes[11]. The BDNF gene has a common single nucleotide polymorphism (SNP). There are genotype variations, where either one or two Met alleles are present. Met allele carriers show deficiencies in activity-dependent release of BDNF, which leads to a downregulation of long-term potentiation and diminished synaptic plasticity in animal models[12]. Human carriers of Met alleles exhibit reduced hippocampal volume and episodic memory storage[13,14], as well as deficits in learning and memory, increased motor impairment post-stroke[15], and worse subcortical stroke outcomes[16]. It is predicted that due to downregulated BDNF secretion, Met allele carriers have more severe post-stroke aphasia compared to homozygotes with Val^66^Val alleles.

There is sparse and inconsistent evidence regarding the effects of BDNF polymorphism on aphasia. de Boer and colleagues[17] tested 53 subjects approximately 1-month post-stroke to examine improvement on communication in daily life situations (Amsterdam-Nijmegen Everyday Language Test[18]) and confrontation naming (Boston Naming Test[19]). Participants were tested pre and post speech-language therapy, which was individualized in an inpatient rehabilitation setting where treatment type and duration varied widely. Subjects showed significant improvement on both language measures. However, there were no group differences based on BDNF genotype and language performance varied at both timepoints regardless of polymorphism. Established prognostic factors like stroke severity and lesion size were not examined in this study, and could have accounted for additional variance in aphasia severity[1]. In contrast, a study by Fridriksson and colleagues[20] reported that Met allele carriers had more severe naming impairments and overall aphasia than Val^66^Val carriers. Notably, these studies’ participants differed in subacute versus chronic stages of aphasia. Thus, these discrepant findings could reflect potential differences in spontaneous recovery associated with BDNF polymorphism. One aim of the current study is to reconcile this inconsistent evidence regarding whether BDNF polymorphism is predictive of aphasia severity following stroke. Moreover, the current study examined whether BDNF accounts for additional variation above and beyond established predictors of aphasia severity, such as lesion size, age at stroke, and cortical excitability. We expected not only that BDNF polymorphism would predict aphasia severity[20], but also that genotypes might interact with other established predictors to provide a more accurate predictive model of aphasia.

Second, we examined ***motor-evoked potentials (MEPs)***, as a neurophysiological indicator of cortical excitability and neuroplasticity. MEPs were measured before and after inhibitory continuous theta burst stimulation (cTBS) – a type of repetitive transcranial magnetic stimulation (rTMS) that requires less than a minute of application to induce comparable neuromodulatory effects[21] and is sensitive to intrinsic differences in cortical excitability and neuroplasticity[22–24]. In particular, we investigated MEP amplitudes at baseline to measure cortical excitability as well as cTBS-induced MEP suppression (difference between pre-vs post-cTBS MEP amplitudes) as a measure of neuroplasticity. Examining MEPs enables us to objectively assess propensity for response to non-invasive brain stimulation (NIBS)[25–28], which is critical to understanding the mechanisms of efficacy and inter-individual variability in NIBS responsiveness for post-stroke language recovery and aphasia rehabilitation.

We explored MEPs as an index of language recovery in aphasia due to evidence that MEPs index domain-general individual differences in cortical excitability and neuroplasticity. Neuroplasticity varies considerably in neurotypical populations, and MEPs are one way to elucidate those differences[29–31]. Accordingly, we hypothesized that MEPs may predict recovery not only from damage to the motor cortex, but from any neural injury. Furthermore, there is a large literature supporting associations between motor and language systems. Lesions to motor cortex have been associated with anomia[32,33] and impaired word comprehension[34,35]. NIBS to pre-motor and motor cortex also implicates the motor strip in speech processing[36,37], phonological and lexical processing[38], and aphasia treatment outcomes[39]. Notably, Glize and colleagues[40] demonstrated that cortical excitability of upper limb motor areas predicted post-stroke language recovery. They found that in addition to aphasia severity and lesion volume, a resting motor threshold (rMT) ratio of the upper limbs measured 1-14 days post-stroke improved abilities to predict language recovery at 6 months post-stroke. This body of evidence indicates that motor cortex plays a role in language, and that the functional integrity and excitability of the corticomotor pathway may be a potential predictor of aphasia severity.

Last, we explored the degree to which ***BDNF polymorphism interacted with other metrics of plasticity*** to improve aphasia severity predictions compared to established predictors. In particular, the metrics of plasticity we examined included cortical excitability, stimulation-induced neuroplasticity, and age at stroke, which is an established predictor of post-stroke severity that is also associated with plasticity[41,42]. Interactions were critical to examine based on evidence that BDNF polymorphism modulates stimulation-induced plasticity[20,43]. Parchure and colleagues[43] examined whether BDNF polymorphism impacts cTBS-induced MEP suppression in chronic stroke patients. They found that Val^66^Val carriers exhibited immediate MEP suppression, which is the expected inhibitory response to cTBS [22,44,45]. In contrast, Val^66^Met carriers exhibited a delayed cTBS response, such that MEPs decreased 30-minutes post-stimulation. This suggests that BDNF polymorphism influences MEP response to neuroplasticity-inducing stimulation protocols, such as cTBS, in chronic stroke patients. However, clinical applications of this finding have not been explored. The current work is, to our knowledge, the first to investigate the utility of genetic and neurophysiological biomarkers of neuroplasticity in predicting post-stroke aphasia severity. We hypothesized that individual variation in MEPs both before and after cTBS may predict severity, and that this effect may furthermore be modulated by BDNF polymorphism.

Indeed, BDNF polymorphism has predicted language outcomes after combined neurostimulation and behavioral aphasia treatment. In the study discussed above, Fridriksson and colleagues[20] found that BDNF polymorphism predicted response to anodal transcranial direct current stimulation (A-tDCS) during aphasia treatment. Sixty-six participants with chronic stroke aphasia were assigned to the A-tDCS or sham condition, followed by a 45-minute behavioral treatment, where participants indicated whether an audiovisual word matched a picture. Proportional changes in correct naming were compared at multiple time points. There was no significant effect of BDNF for the sham group. However, in the A-tDCS group, Val^66^Val carriers showed significantly more proportional change in correct naming compared to subjects with the Met polymorphism. This is consistent with predictions that NIBS enhances aphasia treatment effects by modulating long-term synaptic plasticity, which is a process that relies on BDNF secretion. We predicted the same mechanism may influence aphasia severity, and therefore that BDNF polymorphism may interact with cortical excitability, stimulation-induced neuroplasticity, and age at stroke to predict severity.

To summarize, our central hypothesis was that individual differences in neuroplasticity contribute to language recovery and should improve predictive models of post-stroke aphasia severity in the chronic stage. To evaluate this, we examined three aims. First, we assessed whether a genetic biomarker of neuroplasticity – BDNF polymorphism – improves prognostic estimates of aphasia severity from established predictors derived from a patient’s neurological (lesion size) and demographic (time post-stroke, age at stroke) profile. Second, we assessed whether neurophysiological indicators of neuroplasticity – cortical excitability and stimulation-induced neuroplasticity – improve prognostic estimates of aphasia severity. Third, we explored the degree to which BDNF polymorphism may interact with other metrics of plasticity (cortical excitability, stimulation-induced neuroplasticity, age at stroke) when established predictors like lesion volume and time post-stroke are controlled. Overall aphasia severity, measured by the Western Aphasia Battery Aphasia Quotient (WAB-AQ[46]), served as a metric for the degree to which language functions recovered post-stroke.

## Methods

### Participants

Subjects provided informed consent prior to participation, in accordance with the University of Pennsylvania Institutional Review Board. Participants were 19 subjects with a single left-hemisphere ischemic stroke that occurred >6 months prior to participation. All subjects completed a full dataset, which included BDNF genotyping, MEP, cTBS, and WAB testing. Each procedure is described below. Groups did not significantly differ in age (t=0.69, p=0.50), education (t=0.85, p=0.41), time post-stroke (t=0.35, p=0.73), or lesion volume (t=-1.43, p=0.17). See Table 1 for demographic and neurophysiological data. One participant was excluded from analysis due to performing higher than the recommended clinical cut-off for aphasia diagnosis. This resulted in a final sample of 18 participants.

**Table 1.**
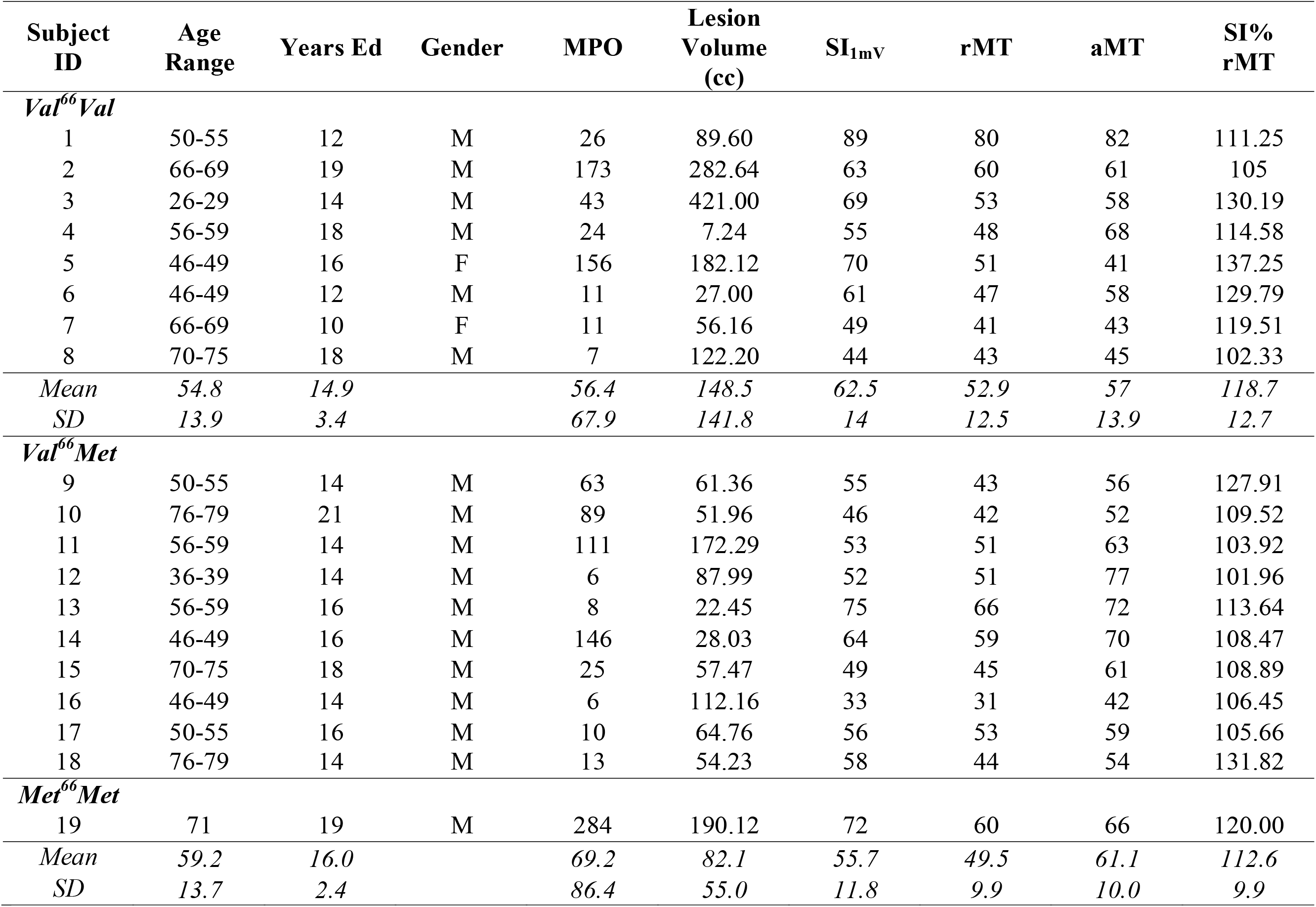
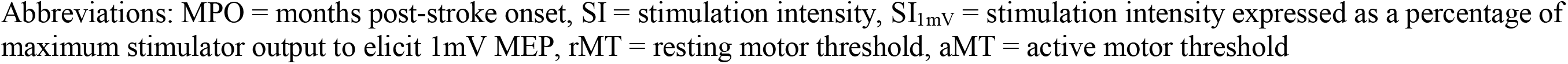
Participant Demographics, Neurophysiological Characteristics, and Stimulation Parameters.

| Subject ID | Age Range | Years Ed | Gender | MPO | Lesion Volume (cc) | SI<sub>1mV</sub> | rMT | aMT | SI% rMT |
| --- | --- | --- | --- | --- | --- | --- | --- | --- | --- |
| *Val<sup>66</sup>Val* |  |  |  |  |  |  |  |  |  |
| 1 | 50-55 | 12 | M | 26 | 89.60 | 89 | 80 | 82 | 111.25 |
| 2 | 66-69 | 19 | M | 173 | 282.64 | 63 | 60 | 61 | 105 |
| 3 | 26-29 | 14 | M | 43 | 421.00 | 69 | 53 | 58 | 130.19 |
| 4 | 56-59 | 18 | M | 24 | 7.24 | 55 | 48 | 68 | 114.58 |
| 5 | 46-49 | 16 | F | 156 | 182.12 | 70 | 51 | 41 | 137.25 |
| 6 | 46-49 | 12 | M | 11 | 27.00 | 61 | 47 | 58 | 129.79 |
| 7 | 66-69 | 10 | F | 11 | 56.16 | 49 | 41 | 43 | 119.51 |
| 8 | 70-75 | 18 | M | 7 | 122.20 | 44 | 43 | 45 | 102.33 |
| *Mean* | *54.8* | *14.9* |  | *56.4* | *148.5* | *62.5* | *52.9* | *57* | *118.7* |
| *SD* | *13.9* | *3.4* |  | *67.9* | *141.8* | *14* | *12.5* | *13.9* | *12.7* |
| *Val<sup>66</sup>Met* |  |  |  |  |  |  |  |  |  |
| 9 | 50-55 | 14 | M | 63 | 61.36 | 55 | 43 | 56 | 127.91 |
| 10 | 76-79 | 21 | M | 89 | 51.96 | 46 | 42 | 52 | 109.52 |
| 11 | 56-59 | 14 | M | 111 | 172.29 | 53 | 51 | 63 | 103.92 |
| 12 | 36-39 | 14 | M | 6 | 87.99 | 52 | 51 | 77 | 101.96 |
| 13 | 56-59 | 16 | M | 8 | 22.45 | 75 | 66 | 72 | 113.64 |
| 14 | 46-49 | 16 | M | 146 | 28.03 | 64 | 59 | 70 | 108.47 |
| 15 | 70-75 | 18 | M | 25 | 57.47 | 49 | 45 | 61 | 108.89 |
| 16 | 46-49 | 14 | M | 6 | 112.16 | 33 | 31 | 42 | 106.45 |
| 17 | 50-55 | 16 | M | 10 | 64.76 | 56 | 53 | 59 | 105.66 |
| 18 | 76-79 | 14 | M | 13 | 54.23 | 58 | 44 | 54 | 131.82 |
| *Met<sup>66</sup>Met* |  |  |  |  |  |  |  |  |  |
| 19 | 71 | 19 | M | 284 | 190.12 | 72 | 60 | 66 | 120.00 |
| *Mean* | *59.2* | *16.0* |  | *69.2* | *82.1* | *55.7* | *49.5* | *61.1* | *112.6* |
| *SD* | *13.7* | *2.4* |  | *86.4* | *55.0* | *11.8* | *9.9* | *10.0* | *9.9* |

Abbreviations: MPO = months post-stroke onset, SI = stimulation intensity, SI<sub>1mV</sub> = stimulation intensity expressed as a percentage of maximum stimulator output to elicit 1mV MEP, rMT = resting motor threshold, aMT = active motor threshold

### Overview of Experiment Design

WAB-AQ[46] was the outcome measure, measuring overall post-stroke aphasia severity. Each session began by determining the participant’s rMT and baseline MEPs. Next, we obtained the active motor threshold (aMT) to determine cTBS intensity. Thirty MEPs were obtained before cTBS (baseline) and 10-minutes post-stimulation. Refer to Figure 1. This analysis was embedded in a larger study of motor physiology, in which MEPs were gathered at multiple timepoints. The current analysis focused on the 10-minute post-cTBS MEPs because this timepoint showed peak stimulation effects[43].

**Figure 1.**
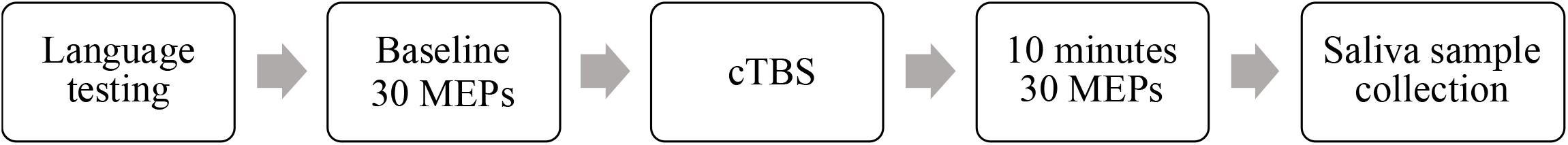
Schematic of Experimental Design. Language testing occurred before cTBS. cTBS was administered for 40 seconds (600 pulses). 30 MEPs were recorded pre-cTBS and at 10 minutes after cTBS. Saliva samples were collected after all stimulation and multiple timepoints of MEP collection were complete.

### BDNF Genotyping

We collected genomic DNA from subject saliva samples, using Oragene® DNA collection kits. Genomic DNA was then isolated using the prepIT.L2Preagent (cat #PT-L2P-5, DNA Genotek Inc, Canada) and precipitated with ethanol according to the manufacturer protocol. The DNA samples were genotyped for BDNF (single nucleotide polymorphism rs6265) using the TaqMan SNP Genotyping Assay (C_11592758_10) designed by Thermo Fisher Scientific. Primers and probes were mixed with TaqMan® Universal PCR Master Mix (Thermo Fisher Scientific). 4.5 μL of genomic DNA (2.5ng/μL) was transferred in triplicate to a 384-well plate, with each well containing 5.5 μL of the PCR mixture. PCR reaction was performed following a protocol provided by ABI. The allele was discriminated by post-PCR plate reading on the ViiA™ 7 System. Genotype data were processed using the ViiA™ 7 Software (Thermo Fisher Scientific).

### Transcranial Magnetic Stimulation (TMS)

We administered single-pulse TMS with a monophasic waveform to the primary motor cortex of the intact right hemisphere using a Magstim 2002 Stimulator with a 70mm figure-eight coil (Magstim Co., Whitland, Dyfield, UK). We uploaded participants’ T1-weighted MRI scans to the Brainsight® Neuronavigation system (Rogue Research, Montreal) to identify the optimal scalp position within the right primary motor cortex for eliciting a MEP from the left first dorsal interosseous (FDI) muscle. In accordance with standard methods, we adjusted the intensity of stimulation such that resulting baseline MEPs for each subject had an average amplitude of ∼1mV[21]. We acquired MEPs as subjects were seated in a chair with their arms resting on their lap or a pillow. Following standard procedures, we defined rMT as the minimum pulse intensity required to elicit MEPs with peak-to-peak amplitudes of at least 50μV in five of ten consecutive trials with the FDI at rest[47,48]. The starting stimulation intensity to acquire MEPs was 110% of rMT, after which the intensity was steadily increased by 1-2% until 10-12 consecutive MEPs were close to 1mV in peak-to-peak amplitude. The coil position was maintained at the optimal scalp location and orientation during the acquisition of MEPs using the neuronavigation system. The single TMS pulses were delivered with an inter-stimulus interval of 6 seconds with a random jitter of 6%. See Table 1 for individual stimulation parameters.

### Electromyography (EMG)

We recorded EMG activity using surface electrodes panning the belly of the FDI muscle of each subject’s left hand, with the ground electrode laced along the left wrist. Signals were amplified and band-pass-filtered between 20-2000 Hz, digitized with a sample-rate of 5 kHz, and stored for off-line analysis using SIGNAL software (Cambridge Electronic Devices, Cambridge, UK).

### Continuous Theta Burst Stimulation (cTBS)

We administered cTBS with a biphasic waveform using a Magstim SuperRapid2 Stimulator (Magstim Co., Whitland, Dyfield, UK). CTBS consisted of a continuous delivery of 50 Hz triplets of TMS pulses at 5 Hz for a total of 600 pulses and approximately 40 seconds. CTBS intensity was 80% of aMT, which is the minimum pulse intensity required to produce MEPs with peak-to-peak amplitudes of at least 200 μV in five of ten consecutive pulses while participants contracted the first dorsal interosseous (FDI) muscle at 20% of the maximum voluntary contraction. To ensure 20% contraction of the FDI, we recorded EMG of each subject’s contracting at maximal force and then practicing at a strength that filled 20% of the maximal EMG bounds. We used the same biphasic stimulator to determine aMT and administer cTBS. All subjects tolerated the cTBS with no adverse effects.

### Statistical Analysis

We used linear regression models in R with aphasia severity (WAB-AQ) as the outcome measure. We used a backward-fitting stepwise regression approach to determine which factors accounted for significant variability in severity. First, we included factors of interest that are common predictors of aphasia, including time post-stroke (log-transformed months post-stroke; LogMPO), age at time of stroke (AgeCVA), and total lesion volume (LesVol). Next, we included predicted indicators of neuroplasticity, including BDNF genotype (Met allele carriers vs. Val^66^Val carriers), baseline MEP (cortical excitability; MEPbase), and cTBS-induced changes in MEP (stimulation-induced neuroplasticity, measured as the difference between pre-and 10-minutes post-cTBS; MEPd10). In addition, we examined potential interactions between BDNF genotype and the following factors: AgeCVA, MEPbase, and MEPd10. Because time-post stroke LogMPO and LesVol were not equally distributed among the different BDNF genotypes in our sample, we controlled for these factors as covariates. The initial model in the stepwise search was as follows:

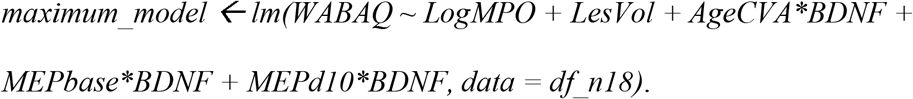

We then used the *step* function of the R *stats* package[49] to implement a backward-fitting stepwise regression approach:

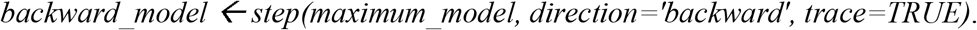

Model comparisons assessed the degree to which each factor and interaction terms significantly improved model fit (Table 2). Factors that did not improve model fit were excluded from subsequent models. We computed variance inflation factor using the R *car* package[50], and found no concerning collinearity between MEP measurements at baseline (VIF=3.53) versus 10 minutes post-stimulation (VIF=2.08).

**Table 2.** Model Comparisons.

|  | Res.Df | RSS | Df | Sum of Sq. | F | Pr(>F) |
| --- | --- | --- | --- | --- | --- | --- |
| *Maximum Model* | 2729 | 170027 |  |  |  |  |
| MEPd10 : BDNF | 2730 | 183707 | -1 | -13680 | 219.57 | <0.001 |
| MEPbase : BDNF | 2730 | 213270 | -1 | -43243 | 694.06 | <0.001 |
| AgeCVA : BDNF | 2730 | 179495 | -1 | -9467.7 | 151.96 | <0.001 |
| BDNF | 2730 | 181988 | -1 | -11961 | 191.97 | <0.001 |
| MEPd10 | 2729 | 170027 | 0 | <0.001 |  |  |
| MEPbase | 2729 | 170027 | 0 | <0.001 |  |  |
| AgeCVA | 2729 | 170027 | 0 | <0.001 |  |  |
| LesVol | 2730 | 205731 | -1 | -35704 | 573.07 | <0.001 |
| LogMPO | 2730 | 170628 | -1 | -600.6 | 9.64 | 0.002 |

Notes: Res.Df: residual degrees of freedom; RSS: residual sum of squares; Df: degrees of freedom; Sum of Sq.: sum of squares. LogMPO: log-transformed months post-stroke; AgeCVA: age at time of stroke/cerebrovascular accident; LesVol: total lesion volume; BDNF: brain-derived neurotrophic factor, including Met allele carriers vs. Val<sup>66</sup>Val carriers; MEPbase: baseline motor-evoked potential (MEP) amplitude, as measure of cortical excitability; MEPd10: difference score between MEP amplitudes from baseline to 10-minutes post-cTBS, as measure of stimulation-induced neuroplasticity.

## Results

### BDNF Genotyping

Among the 18 subjects, 7 were BDNF Val^66^Val carriers, 10 were Val^66^Met allele carriers and 1 was a Met^66^Met carrier. All Met allele carriers were included in a single group because there was an insufficient number of Met^66^Met subjects to analyze separately. Excluding the single Met^66^Met carrier from analyses did not significantly alter the results. Met allele carriers were coded as 0 and Val^66^Val carriers were coded as 1 for our analysis.

### Linear Regression

The following model structure was the result of the backward-fitting stepwise regression:

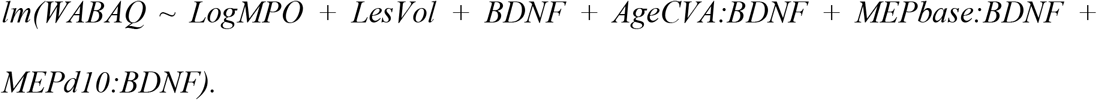

As reported in Table 3, each predictor accounted for significant variance in aphasia severity (adjusted R^2^=0.70). This suggests that when controlling for the effects of time post-stroke (β=-0.63, SE=0.20, t=-3.11, p=0.002) and total lesion volume (β=-0.10, SE=0.004, t=-23.94, p<0.001), BDNF genotype shows a main effect such that when all other factors are constant, Val^66^Val carriers show less severity than Met allele carriers (β=22.68, SE=1.64, t=13.86, p<0.001; Figure 2). Furthermore, BDNF genotype interacted with each predictor of interest: age at stroke, baseline MEP, and change in MEP. Each of these three findings are discussed below, and Table 2 reports pairwise results for each BDNF genotype interaction effect. Model comparisons are reported in Table 3.

**Table 3.**
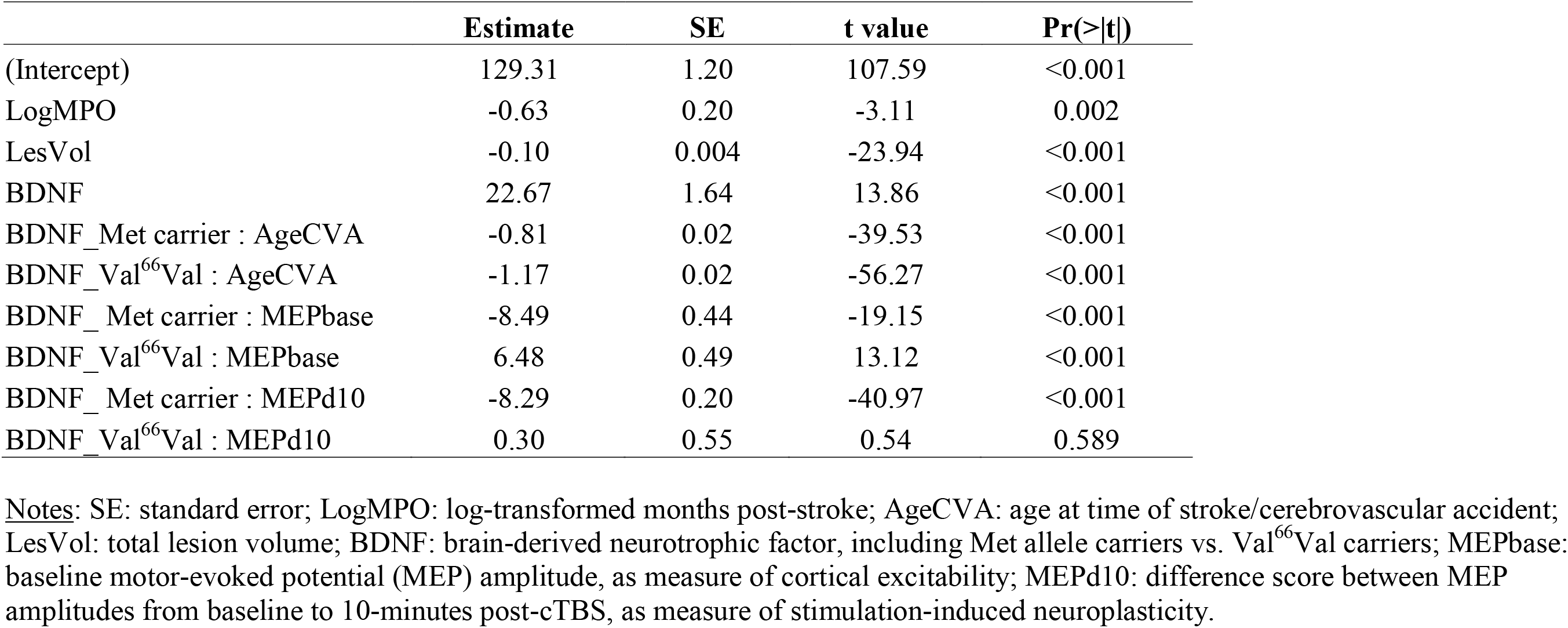
Linear Regression Results.

|  | **Estimate** | **SE** | **t value** | **Pr(>\|t\|)** |
| --- | --- | --- | --- | --- |
| (Intercept) | 129.31 | 1.20 | 107.59 | <0.001 |
| LogMPO | -0.63 | 0.20 | -3.11 | 0.002 |
| LesVol | -0.10 | 0.004 | -23.94 | <0.001 |
| BDNF | 22.67 | 1.64 | 13.86 | <0.001 |
| BDNF_Met carrier : AgeCVA | -0.81 | 0.02 | -39.53 | <0.001 |
| BDNF_Val<sup>66</sup>Val : AgeCVA | -1.17 | 0.02 | -56.27 | <0.001 |
| BDNF_Met carrier : MEPbase | -8.49 | 0.44 | -19.15 | <0.001 |
| BDNF_Val<sup>66</sup>Val : MEPbase | 6.48 | 0.49 | 13.12 | <0.001 |
| BDNF_Met carrier : MEPd10 | -8.29 | 0.20 | -40.97 | <0.001 |
| BDNF_Val<sup>66</sup>Val : MEPd10 | 0.30 | 0.55 | 0.54 | 0.589 |

Notes: SE: standard error; LogMPO: log-transformed months post-stroke; AgeCVA: age at time of stroke/cerebrovascular accident; LesVol: total lesion volume; BDNF: brain-derived neurotrophic factor, including Met allele carriers vs. Val<sup>66</sup>Val carriers; MEPbase: baseline motor-evoked potential (MEP) amplitude, as measure of cortical excitability; MEPd10: difference score between MEP amplitudes from baseline to 10-minutes post-cTBS, as measure of stimulation-induced neuroplasticity.

**Figure 2.**
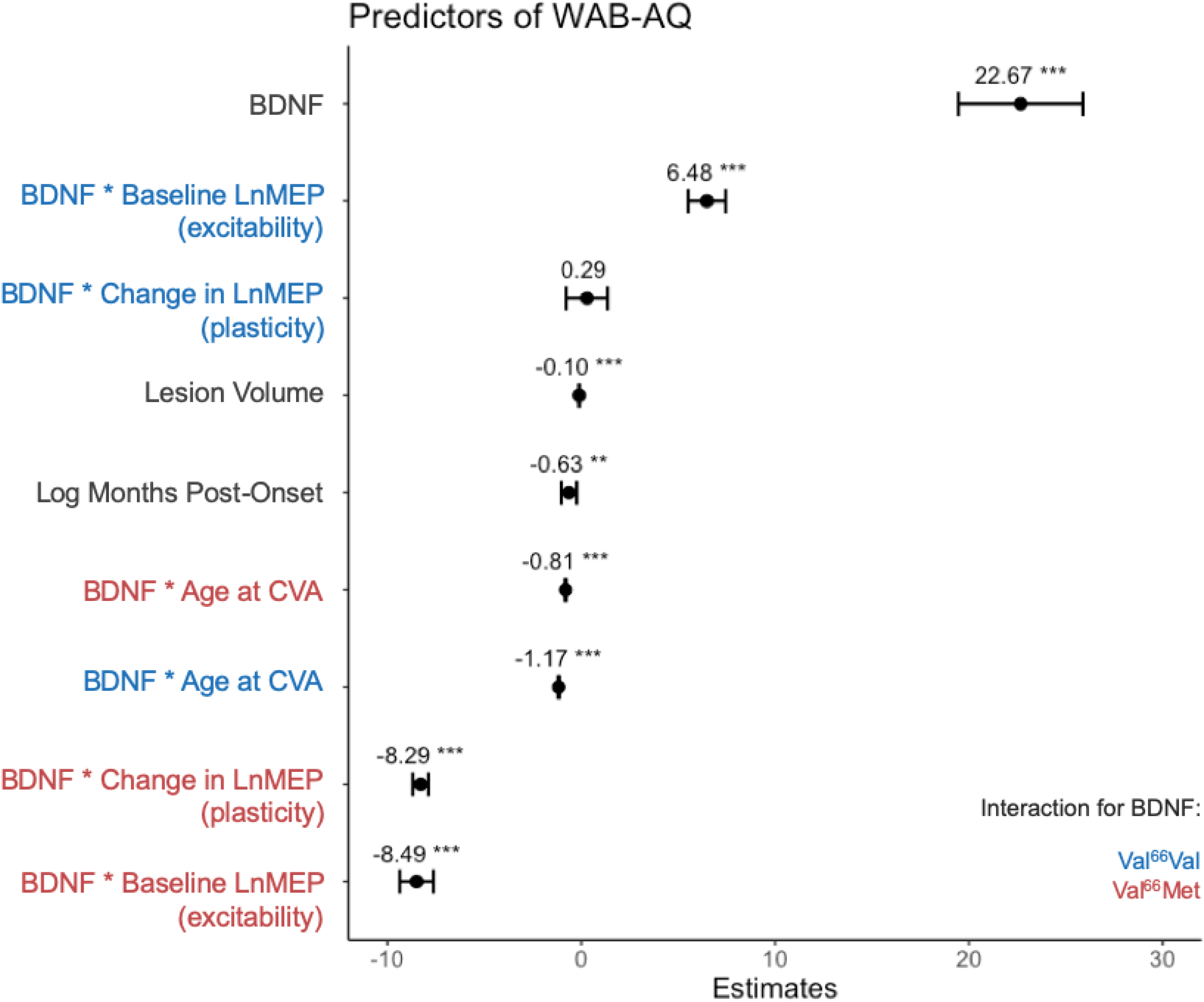
Estimates (Fixed Effects) of Regression Model. <u>Notes</u>: Estimates are beta values, with brackets for standard error. ^**^ denotes significance at p<0.01, ^***^ denotes significance at p<0.001. Val^66^Val carriers are represented in blue and Met allele carriers are in red, interaction effects for each genotype are displayed separately.

#### Age at CVA x BDNF interaction

Increased age at stroke was associated with lower WAB-AQ for both groups, but had a stronger effect on aphasia severity for Val^66^Val carriers (β=-1.17, SE=0.02, t=-56.27, p<0.001) than Met allele carriers (β=-0.81, SE=0.02, t=-39.53, p<0.001; Figure 3A). We conducted post-hoc analyses comparing BDNF genotype groups for the upper and lower terciles of age at CVA in the R package emmeans[51], which revealed that this interaction effect was driven by a significant difference for individuals who were younger at time of CVA. Specifically, Val^66^Val vs. Met allele carriers exhibited less severity at younger (β=-2.99, SE=0.40, t=-7.56, p<0.001) but not older age at CVA (β=-0.003, SE=0.38, t=-0.01, p=1).

**Figure 3.**
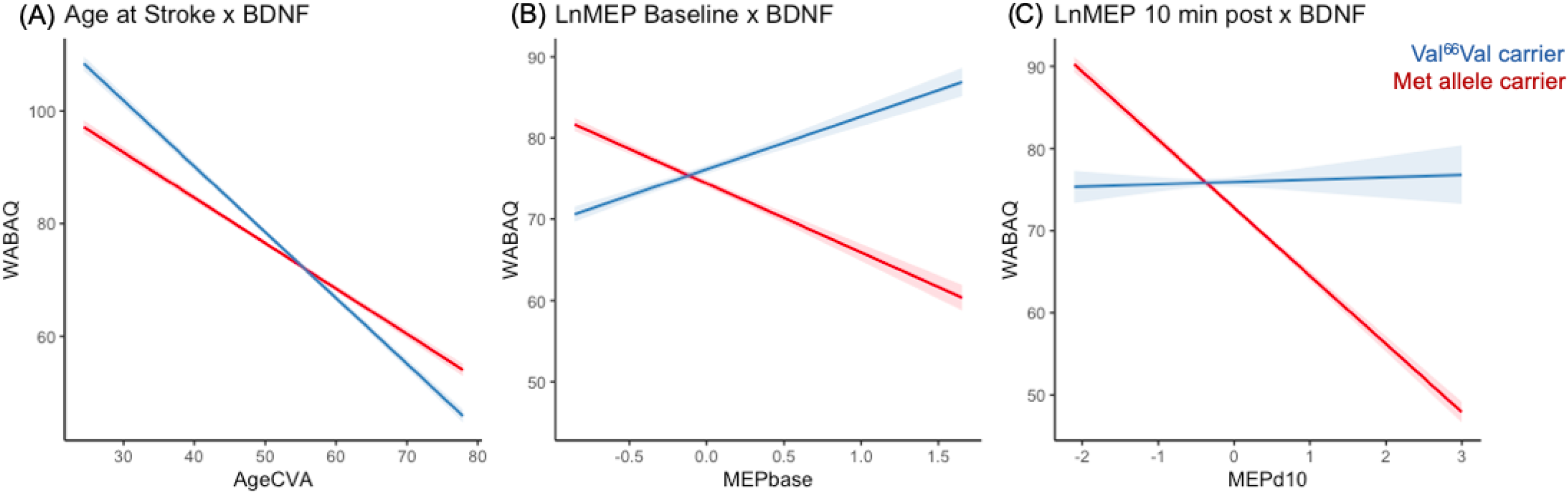
BDNF Interactions Predict WAB-AQ. <u>Notes</u>: X axis is (A) age in years at time of cerebrovascular accident, (B) mean natural log-transformed MEP amplitudes at baseline as an index of cortical excitability, (C) difference between baseline and post-cTBS mean natural log-transformed MEP amplitudes. Y axis (A-C) is predicted WAB-AQ values. Val^66^Val carriers are represented in blue and Met allele carriers are in red.

#### Cortical excitability x BDNF interaction

Cortical excitability was positively associated with WAB-AQ for Val^66^Val carriers (β=6.48, SE=0.49, t=13.12, p<0.001), but negatively associated with WAB-AQ for Met allele carriers (β=-8.49, SE=0.44, t=-19.15, p<0.001; Figure 3B).

#### Stimulation-induced neuroplasticity x BDNF interaction

Worse severity for Met allele carriers (i.e., lower WAB-AQ) was associated with cTBS aftereffects in the opposite direction (i.e., MEP facilitation) (β=-8.29, SE=0.20, t=-40.97, p<0.001), while variability in severity for Val^66^Val carriers was not associated with cTBS aftereffects (β=0.30, SE=0.55, t=0.54, p=0.59; Figure 3C). To better understand the relationship between the latter two findings, we ran correlations between mean MEP at baseline and change in MEP post-10 minutes for each BDNF genotype group. For Met allele carriers, increased excitability (higher baseline MEPs) was associated with decreased MEP suppression (less neuroplasticity; R=0.50, p<0.001). However, for Val^66^Val carriers, increased excitability was associated with greater neuroplasticity (R=-0.26, p<0.001).

## Discussion

This study provides evidence that individual differences in neuroplasticity contribute to aphasia severity. In a sample of participants with chronic post-stroke aphasia, we replicated main effects of established aphasia predictors, including lesion volume and time post-stroke. Moreover, we found that BDNF polymorphism, a genetic biomarker of neuroplasticity, accounted for a substantial amount of variance in aphasia severity and improved estimates of aphasia severity from established predictors alone. In addition, BDNF polymorphism interacted with other metrics of plasticity, including age at stroke[41,42], cortical excitability[29,30], and stimulation-induced neuroplasticity[27,28] to predict aphasia severity. These findings and their implications are discussed below.

The current results indicate that BDNF polymorphism is a critical predictor of variance in aphasia, beyond established predictors. We found that when controlling for the effects of total lesion volume and time post-stroke, Val^66^Val carriers showed less severe aphasia than Met allele carriers. This is in line with Fridriksson and colleagues’[20] findings that Met allele carriers presented with more severe chronic aphasia than Val^66^Val carriers. Although there are also reports that BDNF polymorphism did not influence post-stroke aphasia recovery, these studies differed in critical ways including recovery stage and outcome measures. As discussed in the introduction, De Boer and colleagues[17] examined patients with acute stroke and observed no differences in everyday language or confrontation naming abilities based on BDNF polymorphism. In addition, they did not control for or examine potential effects of critical factors like aphasia severity and lesion size. Notably, our findings are consistent with our mechanistic hypothesis: because Val^66^Val carriers show stronger propensity for neuroplastic change than Met allele carriers[12,44], they will be more likely to recover and present with mild aphasia. Our results are also consistent with a meta-analysis and multiple independent reports that BDNF polymorphism influenced general stroke recovery[52], as well as post-stroke motor[53] and functional recovery[54], such that Met allele carriers demonstrate worse rehabilitation outcomes than Val^66^Val carriers.

Furthermore, BDNF polymorphism interacted with other indicators of neuroplasticity to improve predictions of aphasia severity. First, BDNF polymorphism interacted with age at the time of stroke. Older participants had more severe aphasia for both BDNF genotypes, but Met allele carriers below the age of 50 exhibited worse outcomes when compared to their homozygous Val^66^Val counterparts. This finding again supports predictions that individuals with stronger, more intact neuroplasticity (Val^66^Val carriers and younger individuals) will be more likely to recover and present with mild aphasia in chronic stroke. Furthermore, these results are consistent with findings that Met allele carriers show greater adverse effects of age on performance than Val^66^Val carriers in non-linguistic cognitive domains, as measured by subjective memory complaints, associative and episodic memory performance, and response to increased executive demands[55,56]. This indicates that genetic effects may have a stronger influence on cognition when neural resources are reduced, as in old age[56].

Second, BDNF polymorphism interacted with two MEP indicators of neuroplasticity: cortical excitability and stimulation-induced neuroplasticity. Both indicators of neuroplasticity were particularly informative of aphasia severity for Met allele carriers. We observed distinct patterns for cortical excitability: Greater baseline excitability was associated with less severity for Val^66^Val carriers, while greater cortical excitability was associated with greater aphasia severity for Met allele carriers. Notably, evidence from neurologically healthy adults also shows strong interactions between BDNF polymorphism and baseline MEPs even when stimulation intensity is individually adjusted to minimize differences in baseline MEP amplitudes (e.g., 1mV)[24]. In addition, greater stimulation-induced neuroplasticity (measured by cTBS after-effects of MEP suppression) was associated with greater aphasia severity for Met allele carriers, and the degree to which individuals showed typical MEP suppression correlated with their potential for recovery. In contrast, stimulation-induced neuroplasticity was not associated with aphasia severity for Val^66^Val carriers. To coalesce these findings, we ran post-hoc correlations, which revealed that higher cortical excitability was associated with greater stimulation-induced neuroplasticity for Val^66^Val carriers, whereas higher excitability was associated with less neuroplasticity for Met allele carriers. For Val^66^Val carriers, higher baseline excitability may provide more potential for MEP suppression, similar to neurologically healthy response[21], while more excitable Met allele carriers may instead be less likely to show expected response to neurostimulation[43]. Further work may explore the mechanisms driving this effect, such as potential associations between cortical excitability and delayed MEP suppression following neurostimulation in Met allele carriers.

These findings provide evidence that BDNF polymorphism contributes to individual differences in cortical excitability and stimulation-induced neuroplasticity, indicating a prognostic utility of previous findings that BDNF polymorphism modulates individual responsiveness to stimulation[20,43]. Furthermore, this study showcases how TMS can be used to measure neuroplasticity by leveraging neurophysiological indices of plasticity that inform functional prognostics. MEPs are informative neurophysiological indicators of plasticity that may reflect domain-general individual differences in cortical excitability and plasticity. Future work may explore the utility of these indicators in relation to domains of non-motor functional recovery following neural injury.

A potential limitation of this study is the modest sample size used to examine the behavioral effects of a genetic polymorphism. However, our sample size is comparable to other studies investigating BDNF and/or MEPs in aphasia[43,57,58]. Despite our modest sample, our results show large effect sizes of BDNF polymorphism and robust effects of multiple factors that account for separate variance in aphasia severity. In addition, it remains unclear whether BDNF polymorphism is only a significant predictor of chronic but not acute post-stroke aphasia. Additional work is required to characterize potential differences in spontaneous recovery associated with BDNF polymorphism. Such investigations could elucidate differences between studies finding no effects of BDNF polymorphism in acute aphasia[17] versus robust effects of BDNF in chronic recovery stages[20]. Likewise, neurophysiological responses such as MEP-suppression following NIBS may differ in acute to chronic stages of stroke recovery. Longitudinal investigations may help determine the optimal prognostic time at which biomarkers of neuroplasticity have the strongest predictive power of long-term functional outcomes. Additional predictors, such as stroke location, may further interact with neuroplastic indicators to contribute to the prognostic strength of the current approach[59]. Finally, future work should seek to replicate the current findings in larger samples of stroke patients with and without aphasia.

## Conclusion

BDNF polymorphism is a substantial predictor of chronic post-stroke aphasia severity, above and beyond established predictors. Furthermore, BDNF polymorphism interacted with other indices of neuroplasticity to predict aphasia severity. In particular, BDNF polymorphism interacted with age at stroke, cortical excitability, and stimulation-induced neuroplasticity to improve ability to predict aphasia severity from established descriptive factors. This study is among the first to demonstrate how including biomarkers and neurophysiological indicators of neuroplasticity and cortical excitability significantly improves ability to predict aphasia severity. These findings provide novel insights into potential sources of variability in stroke recovery that may improve aphasia prognostics.

## Data Availability

Please contact the authors for additional information on the data referred to in this manuscript.

